# Entrainment of cortical gamma oscillations predicts improved bradykinesia and dyskinesia in Parkinson’s disease

**DOI:** 10.64898/2026.06.10.26354720

**Authors:** Maria Shcherbakova, Stephanie Cernera, Amelia G. Hahn, Simon Little, Philip A. Starr

**Affiliations:** Department of Neurological Surgery, University of California San Francisco, San Francisco, CA, USA; Department of Neurology, University of Pennsylvania, Philadelphia, PA, USA; Center for Neuroengineering and Therapeutics, University of Pennsylvania, Philadelphia, PA, USA; Department of Neurology, University of California San Francisco, San Francisco, CA, USA; Stritch School of Medicine. Loyola University Chicago. Chicago, IL

## Abstract

**Background:** Deep brain stimulation (DBS) of the subthalamic nucleus (STN) is hypothesized to improve motor symptoms in Parkinson’s disease (PD) by suppressing pathologically elevated beta activity and promoting “prokinetic” gamma activity in the cortico-basal ganglia-thalamo-cortical loop. Advances in bidirectional DBS devices have revealed that stimulation can modify gamma oscillations via subharmonic entrainment, though entrainment’s therapeutic role remains unclear.

**Objectives:** To identify stimulation parameters that entrain motor cortical and STN gamma oscillations in PD at rest and during movement, and examine their association with motor function.

**Methods:** Sensorimotor cortex and STN field potentials were collected using a bidirectional DBS system in four subjects with PD over a range of stimulation amplitudes and frequencies. Entrainment amplitude at half the stimulation frequency was quantified at rest and during a finger-tapping task in the ON-medication state. The presence or absence of entrainment was studied as a physiomarker of motor symptom severity.

**Results:** The amplitude of stimulation-entrained gamma oscillations was non-linearly related to stimulation intensity and frequency and varied by stimulation contact choice. Entrainment amplitude was highest in precentral gyrus and increased with movement. In the ON-medication state, precentral gyrus gamma entrainment was associated with reduced bradykinesia, dyskinesia, and dystonia. Subthalamic gamma entrainment predicted improved dystonia but was a less significant marker for motor benefit than cortical entrainment.

**Conclusions:** Stimulation-entrained gamma oscillations in the motor network are a physiomarker for optimal DBS response in PD, and could have a role in physiology-guided DBS programming, complementing existing strategies based on suppression of basal ganglia beta activity.

## Introduction

Deep brain stimulation (DBS) of the subthalamic nucleus (STN) is a well-established neurosurgical treatment of Parkinson’s disease (PD) that reduces motor symptoms, and allows reduction of antiparkinsonian medication dose.^1^ DBS is suggested to provide therapeutic benefit by disrupting pathological activity in the basal ganglia-thalamocortical network.^2^ However, its mechanism is incompletely understood. Recently, brain recordings from bidirectional DBS devices — a novel class of pulse generators capable of simultaneous neurostimulation and sensing — have enabled the search for neural signatures of motor sign severity and stimulation treatment responses in patients undergoing chronic neurostimulation.^3^

In PD, abnormal oscillatory activity in the basal ganglia and cortex is linked to movement dysregulation, with beta oscillations (13-30 Hz) considered *antikinetic* due to the association of exaggerated cortico-basal ganglia beta synchrony with bradykinesia. Conversely, finely tuned gamma (FTG; also called “narrowband” gamma) oscillations at 60-90 Hz are considered *prokinetic*, associated with improved OFF-medication motor signs.^4^ Most studies of oscillatory phenomena in PD, and their use in guiding DBS programming, have focused on beta band activity. Yet, all major therapeutic interventions for PD (levodopa,^4^ DBS^5,6^ and basal ganglia lesioning^7^) induce FTG in the motor network, suggesting a relationship to therapeutic mechanism across modalities.

During therapeutic DBS, the peak frequency of FTG may shift — or “entrain” — to a subharmonic of stimulation frequency, usually at half the stimulation frequency (1:2 entrainment).^8^ Stimulation-entrained gamma activity has been proposed as a reliable marker of the ON-medication state,^9,10^ a possible signature of improved motor function,^9^ and a promising control signal for adaptive DBS.^11^ Entrainment phenomena in the motor network are common and occur in response to stimulation at both STN and pallidal targets in several movement disorders.^9,12,13^ However, limited understanding of the stimulation parameters that optimize entrainment, and of its effects on parkinsonian motor signs, impedes clinical translation. Here, we characterize entrained gamma oscillations in the motor network in PD under controlled in-clinic conditions using an investigational sensing-enabled neurostimulator connected to both subthalamic and sensorimotor cortical electrodes. We show that entrained gamma activity is influenced by specific stimulation frequency-amplitude combinations, by movement, and by recording site (STN or cortical). Entrainment was associated with reduced bradykinesia and dyskinesia. The findings suggest a novel paradigm for neurostimulator programming based on selecting parameters that efficiently entrain gamma oscillations.

## Methods

### Subject recruitment and device implantation

Subjects (Table 1) were recruited from a pool of 17 individuals with PD participating in a parent study of chronic brain recording and adaptive DBS at the University of California, San Francisco (ClinicalTrials.gov ID NCT03582891; Institutional Review Board number 18-24454). Subjects in this cohort have provided data to prior publications focusing on adaptive deep brain stimulation^11,14^ and chronic ambulatory brain recording.^12,15,16^ Subjects were bilaterally implanted with an investigational DBS device (Medtronic™ Summit RC+S model B35300R) connected to a quadripolar STN-DBS lead (Medtronic™ model 3389) and to a quadripolar electrocorticography (ECoG) paddle (Medtronic model 0913025) placed subdurally over the sensorimotor cortex (Fig. 1A, B). Criteria for implantation, surgical procedure, and RC+S technical details have been previously described.^11,16^ Further details on selection of subjects and recording hemisphere are in Supplemental Methods.

**Table 1.** Patient demographics.

| Hemisphere | Clinical stimulation settings | Duration of DBS therapy at data collection | Additional motor signs (bradykinesia + ) | LEDD (mg) | Range of settings tested |  |  |
| --- | --- | --- | --- | --- | --- | --- | --- |
|  |  |  |  |  | Contact * | Amplitude (mA) | Frequency (Hz) |
| RCS05 R ** | 3.9 mA, 60 us, 130 Hz | 4 years 2 months | dyskinesia, tremor | 450 | 1 | 0.5 - 5.0 | 90 - 170 |
| RCS17 L | 2.3 mA, 60 us, 130 Hz | 12 months | dyskinesia | 535 | 2 | 0.5 - 4.0 | 90 - 180 |
| RCS18 L | 1.8 mA, 60 us, 130 Hz | 9 months | N/A | 1490 | 0-3 | 1.8 - 5.0 | 40 - 160 |
| RCS18 R | 3.1 mA, 60 us, 130 Hz | 9 months | dystonia | 750 *** | 2 | 0.5 - 5.0 | 50 - 170 |
| RCS20 L | 4.0 mA, 90 us, 130 Hz | 12 months | dyskinesia, tremor | 551 | 2 | 0.5 - 6.0 | 50 - 150 |

\* Stimulating contacts are labeled 0 to 3, from ventral to dorsal. For all hemispheres, tested contact matched that used for clinical stimulation except for RCS18 L (clinical contact = 2).
\*\* Subject RCS05 only contributed cortical data to the study.
\*\*\* RCS18 leads were implanted in a staged fashion, with the left hemisphere placed first and the right hemisphere implanted 3 months later. Data were collected 9 months after each implantation (therefore 3 months apart). Antiparkinsonian medications were reduced between the left- and right-sided assessments to reduce dyskinesia.

**Figure 1.**
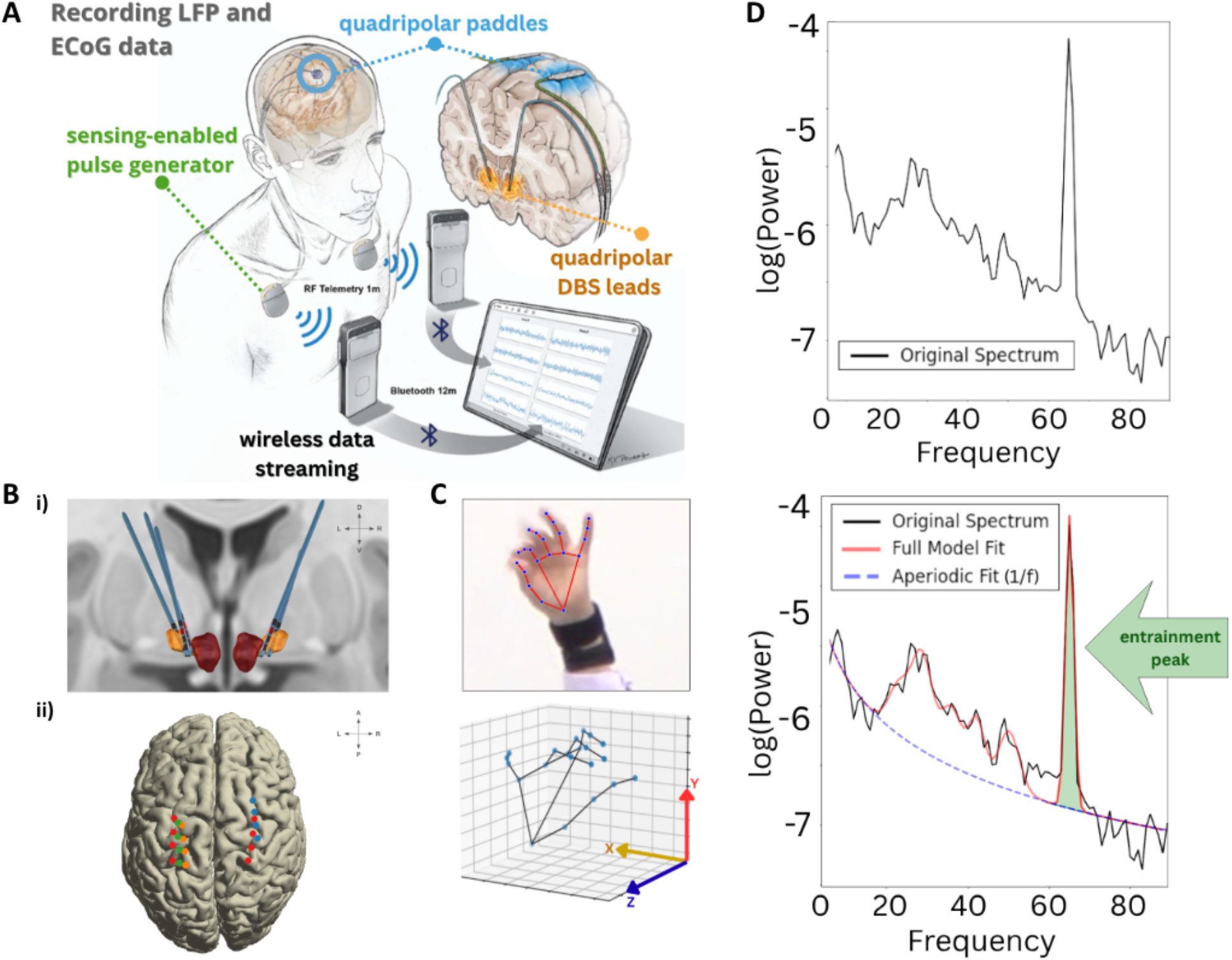
Subject data streaming setup, electrode localization, and experimental paradigm. **A:** Subthalamic and cortical neural recordings were obtained using a chronically implanted investigational bidirectional neural interface, Medtronic Summit RC+S, simultaneously with active STN DBS. All data were wirelessly transmitted to and securely stored on a tablet PC. **B *(across subjects)*: (i)** Localization of quadripolar sensing-enabled DBS leads in the STN, contacts numbered 0 (most ventral) to 3 (most dorsal); stimulating contact is highlighted in red. **(ii)** Localization of quadripolar electrocorticography paddles implanted over somatosensory (S1) and motor (M1) cortices, with contacts numbered 8 (most posterior) to 11 (most anterior); each electrode color represents placement in a specific patient. **C:** Automated motion capture system for bradykinesia assessment. Tapping symptoms were quantified frame by frame from the relative positions of the 21 inferred three-dimensional landmarks of a hand (blue dots). **D:** We used the FOOOF algorithm to quantify entrainment amplitude across a range of stimulation amplitude-frequency pairs. FOOOF extracts an aperiodic component and a number of periodic components to identify peaks in the power spectrum. In this example, entrained gamma activity is present at 65 Hz (half the active stimulation frequency, 130 Hz).

### Experimental design

All subjects’ recordings were conducted in-clinic in the ON-medication state. Each session began with a stimulation washout period of 10 minutes. The high dopaminergic state was confirmed by the presence of cortical levodopa-induced FTG with stimulation off. We performed frequency-amplitude titrations unilaterally, stimulating the STN from a single electrode contact on the DBS lead in increments of 0.5 mA. The parameter ranges varied between hemispheres and subjects due to differences in tolerability. STN and motor cortical field potentials ipsilateral to the stimulation site were concurrently recorded, with contralateral stimulation disabled to avoid cross-hemisphere effects of stimulation. At every frequency-amplitude combination, subjects were instructed to remain at rest for 30 seconds, then to complete a finger-tapping task contralateral to the active stimulation site for 15 seconds.

To assess the relationship between gamma entrainment and motor function, all sessions were video-taped and assessed offline by a blinded rater for presence or absence of dyskinesia, dystonia, and tremor during both rest and movement. Finger-tapping kinematics were extracted from video for quantitative analysis using customized hand-tracking software (Fig. 1C, details in Supplemental Methods). Per MDS-UPDRS convention,^17^ only the first 10 taps were considered for analysis. For each tapping cycle, we computed the mean, maximum, and temporal decrement for tap velocity and tap distance.

### Neural signal collection

In the STN, local field potentials were recorded in a bipolar configuration from two contacts adjacent to the stimulating cathode (Fig. 1B (i)). Two sensorimotor cortical montages were obtained using non-overlapping electrode pairs covering precentral and postcentral gyri (Fig. 1B (ii)). Electrode positions were confirmed using the Lead-DBS toolbox, as detailed previously.^11^ Neural data were sampled at 250-500 Hz, wirelessly streamed to an external tablet computer, and securely stored for post-hoc processing (Fig. 1A).

### Signal processing

We quantified entrainment at each stimulator setting and movement condition using MATLAB 2024a (MathWorks). Epochs of neural data were segmented into 1-second intervals. Epochs with packet loss were removed. Power spectral density (PSD), spanning 1 Hz to the Nyquist frequency with 1-Hz resolution, was estimated using Welch’s method (*pwelch*; 1-second window, 0% overlap to accommodate non-consecutive data packets). Spectra were flattened and the aperiodic component removed (Fig. 1D).^18^ Entrainment was considered present when the spectral peak centered at half stimulation frequency exceeded two standard deviations above the aperiodic-removed spectrum. In trials with entrainment, entrainment amplitude was defined as its log power above the aperiodic baseline. For each stimulation frequency-amplitude combination, entrainment amplitude was normalized across all recording sites within each subject to a common scale ranging from 0 to 10 to allow for consistent comparison.

### Statistics

We evaluated the effect of the recording site and movement state on entrainment amplitude using Type III ANOVA and Tukey’s test. The association between subthalamic and cortical entrainment was measured with the Chi-squared statistic and Cramer’s *V*. We evaluated clinical effects of entrainment by using two complementary approaches. First, we used the Chi-squared statistic, to relate the presence or absence of entrainment to the presence or absence of motor signs. Second, we fit linear mixed-effects models, to assess how the presence or absence of entrainment affected each video-derived kinematic metric (see Experimental design), with outcomes expressed as within-subject z-scores. DBS amplitude was incorporated as a fixed-effect covariate to dissociate effects of entrainment from those related to stimulation amplitude, and hemispheres were modeled as random intercepts.

## Results

While off stimulation and ON-medication, all five hemispheres exhibited levodopa-induced FTG in the 79-90 Hz range in the cortex and subthalamic nucleus (example in Figure 2A). When stimulation produced entrainment, this generally occurred at half the stimulation frequency (the second subharmonic; Fig. 2A). Thus, only 1:2 entrainment was further quantified.

**Figure 2.**
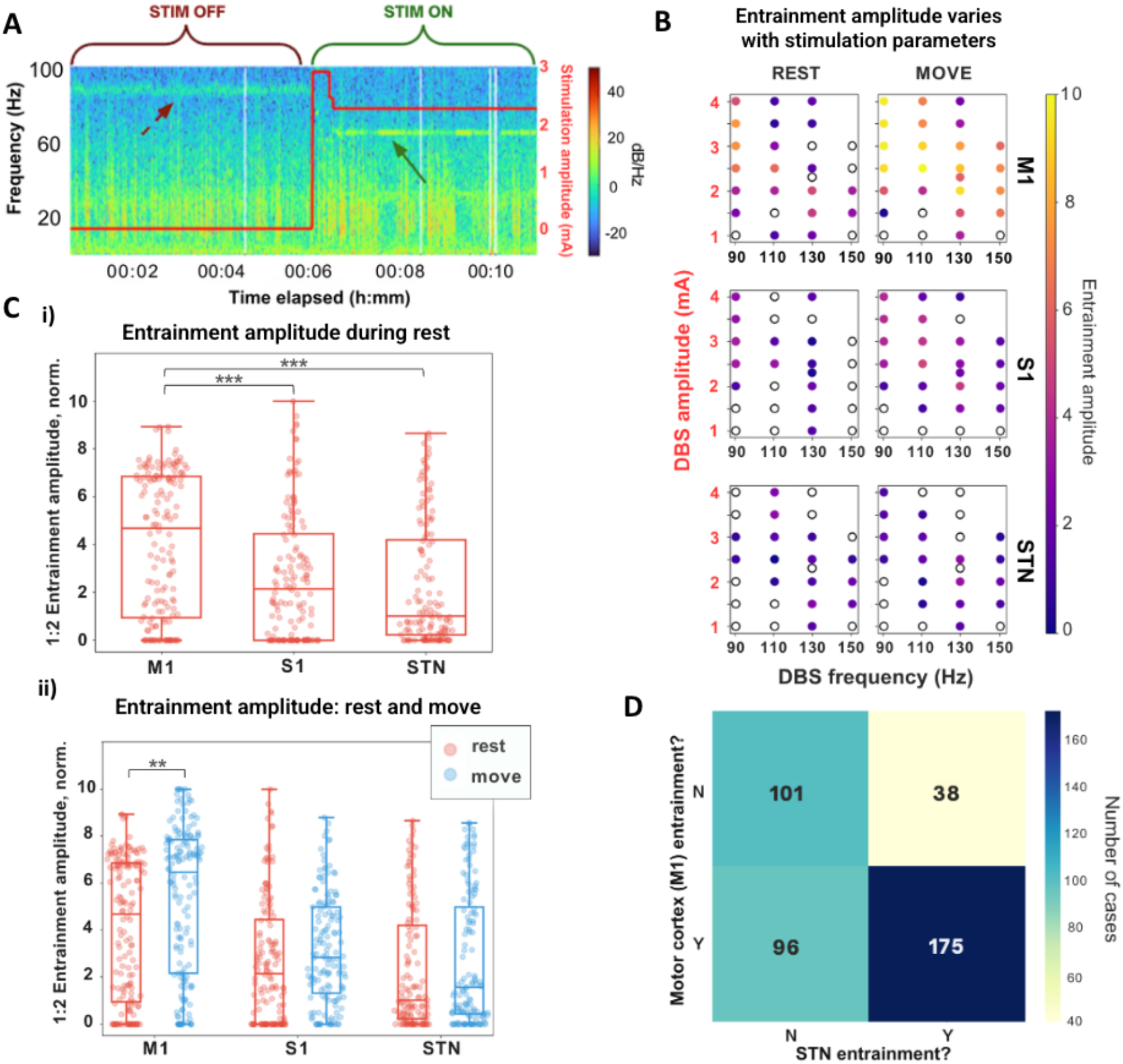
Effects of stimulation parameters, recording site, and movement on entrainment. **A:** An example of cortical gamma activity recorded from the anterior electrode pair (precentral gyrus) in the ON-medication state, ON and OFF stimulation (red line indicates stimulation amplitude). At 2.3 mA, levodopa-induced gamma oscillations that were present at 80-90 Hz in the absence of DBS now entrain to 65 Hz (the half harmonic of stimulation frequency). **B:** A single hemisphere example of the effects of different stimulation frequency-amplitude combinations on the amplitude of entrained gamma activity at half stimulation frequency, at precentral gyrus, post-central gyrus, and subthalamic recording sites during rest and movement. Each dot represents one frequency-amplitude combination. Empty circles indicate trials where no entrainment peak above aperiodic was detected. **C (i), (ii):** Boxplots of entrained amplitudes across all subjects and all stimulation parameters, comparing effects of recording site and movement state. Entrainment had the highest amplitude in the precentral gyrus during both rest and movement conditions (*p*<0.001). Movement (finger tapping) produced a significant increase in entrainment amplitude in M1 (*p*<0.01) only. *(Each dot represents a single frequency-amplitude combination trial, normalized across recording sites per subject; trials that resulted in entrainment in neither movement state were excluded)*. **D:** 2x2 contingency table for presence and absence of motor cortex and STN entrainment. 1:2 entrainment was detected concurrently in the STN and M1 in 57% of all entrainment-positive trials across subjects and movement states. While instances of STN and M1 entrainment were found to be moderately associated (*χ*^*2*^ = 49.6 (*p*<0.001); Cramér’s *V* = 0.353), 35% of trials in which motor cortical entrainment occurred, were not accompanied by simultaneous STN entrainment.

### Entrainment is related to stimulation parameters, recording site, and movement state

For each hemisphere, the presence and the amplitude of entrained activity were scored across a wide range of stimulation frequencies and amplitudes, both at rest and during the movement task. Entrainment was scored at three recording sites: precentral gyrus, postcentral gyrus, and subthalamic nucleus. A heatmap of entrainment amplitudes for all tested stimulation parameters is shown for a representative hemisphere in Figure 2B, and for all hemispheres in Figure S1.

Entrained activity was observed across recording sites in all hemispheres, but the exact entrainment-inducing set of stimulation settings was specific to each hemisphere (Fig. 2B, S1). In general, entrainment was not monotonically related to stimulation amplitude as increases in stimulation amplitude could result in weakening or loss of entrainment, as predicted by modelling studies^19^ (Fig. S1; e.g., see RCS17 at 130 Hz). Previous modelling studies^19^ predicted an asymmetry (“left lean”) in the region of parameter space where entrainment occurs, specifically a pattern in which there is a steeper cutoff of entrainment at high frequencies than low frequencies. This pattern is clearly seen for hemispheres RCS18R and RCS20L, in the two subjects who tolerated the most extensive exploration of the stimulation parameter space (Fig. S1). Although we focused primarily on typical clinical stimulation frequencies (100-180 Hz), whose half-harmonic entrainment was in the gamma range, in three hemispheres we also examined entrainment at low stimulation frequencies (≤60 Hz). Half-harmonic beta entrainment occurred at all sites tested (Fig. S1).

We found a main effect of both the recording site (*F* = 55.08, *p* < 0.001, *df* = 2; Fig. 2C) and movement state (*F* = 11.40, *p* < 0.001, *df* = 1) on entrainment amplitude. The interaction terms were insignificant (*F* = 2.02, *p* = 0.1, *df* = 2). Separate Tukey’s tests for each variable revealed that entrainment had the highest amplitude in the precentral gyrus during both rest and move conditions (*p* < 0.001), and that the precentral gyrus exhibited the largest movement-related amplitude increase (*p* = 0.003), compared to the postcentral gyrus (*p* = 0.14) and STN (*p* = 0.28).

Of note, motor cortical and subthalamic entrainment did not always co-occur (Fig. 2D). Approximately a third of trials with motor cortical entrainment did not have co-occurring subthalamic entrainment, while 18% of trials with STN entrainment lacked motor cortex entrainment.

### Precentral gyrus entrained gamma is strongest when using the stimulation contact within dorsal STN

In general, entrainment was studied only during stimulation at the STN contact utilized for chronic therapy (Table 1), as study across all contacts would have placed an excessive burden on most subjects. In one hemisphere (RCS18 L), however, we were able to evaluate the effect of stimulation contact choice across all four contacts. The contact used clinically for chronic therapy (contact 2, dorsolateral STN) resulted in most prominent entrainment across a range of DBS amplitudes (Fig. 3).

**Figure 3.**
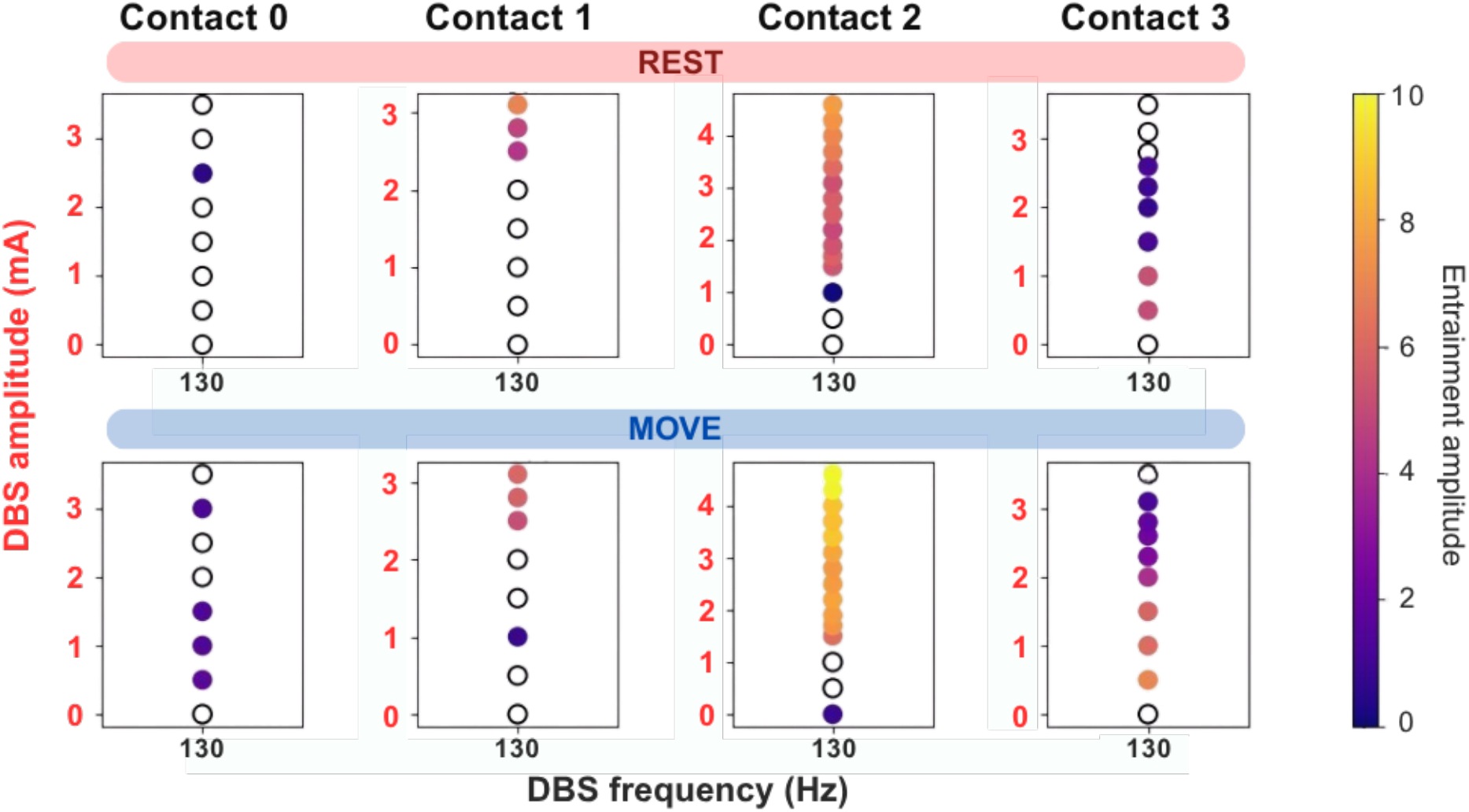
Effects of stimulation contact choice on cortical gamma entrainment at 130 Hz. We assessed the amplitude of entrained gamma recorded from the anterior cortical electrode pair while varying STN stimulation amplitude at each electrode contact individually, from the most ventral (Contact 0) to the most dorsal (Contact 3), at a constant frequency of 130 Hz (the frequency used clinically) in a single hemisphere. STN contact choice influences the amplitude of entrained oscillations in the motor cortex, with contact 2 stimulation (dorsal STN and also the contact used clinically) resulting in highest entrainment amplitudes.

### Presence of motor cortical entrainment is associated with improved motor signs

For all subjects and stimulation parameters, we related the presence or absence of gamma entrainment with the presence or absence of specific motor signs: dyskinesia (present in *n*=3 subjects), dystonia (*n*=1 subject), and tremor (*n*=2 subjects) (Fig. 4A). Both motor cortical and STN entrainment predicted the absence of dystonia (M1: *χ*^*2*^ = 6.57, *p* = 0.010; STN: *χ*^*2*^ = 8.25, *p* = 0.004). Motor cortical, but not STN, entrainment predicted the *absence* of dyskinesia (M1: *χ*^*2*^ = 5.23, *p* = 0.022; STN: *χ*^*2*^ = 0.139, *p* = 0.71). We did not find a relationship between entrainment and tremor (M1: *χ*^*2*^ = 0.097, *p* = 0.76; STN: *χ*^*2*^ = 0.103, *p* = 0.75).

**Figure 4.**
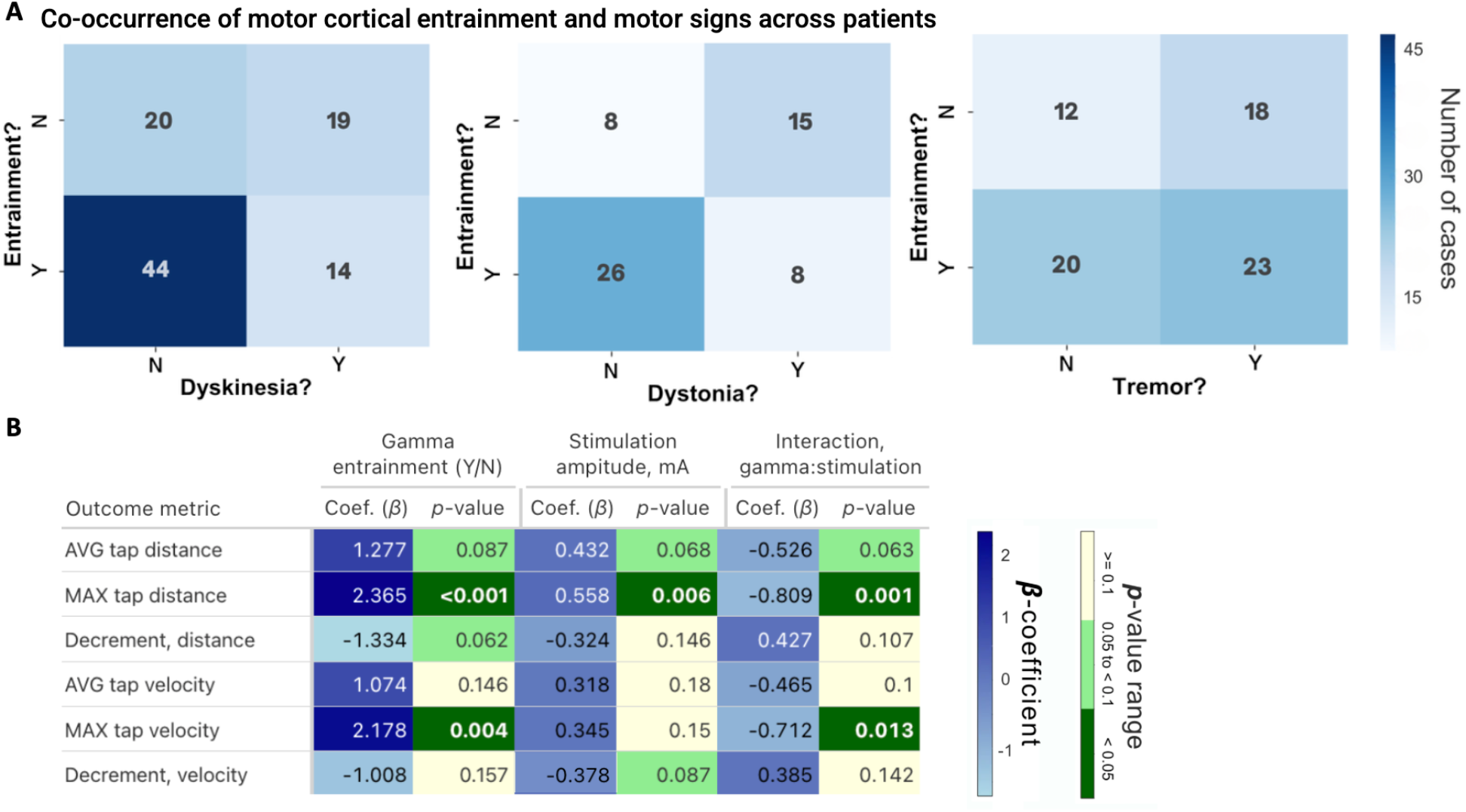
Effects of motor cortical gamma entrainment on motor function. **A**: 2x2 contingency tables showing the association between motor cortical gamma entrainment and common parkinsonian motor signs observed in clinic. **B:** Summary of the results of a linear mixed effects model to quantify the effects of gamma entrainment at stimulation frequencies ≥100 Hz on predefined metrics of bradykinesia obtained from video kinematics. As predictors, the model uses 1) presence of cortical gamma entrainment, recorded immediately *prior* to movement onset (finger-tapping task), and 2) stimulation amplitude. *Coefficients show the direction and magnitude of relationships, with greater absolute β-coefficient values signifying stronger associations*.

In four out of five hemispheres, we assessed the effect of gamma entrainment on bradykinesia from finger-tapping kinematics *(RCS20 L was excluded from this analysis, as video footage was unavailable for stimulation frequencies* ≥*100Hz)*. We measured tapping distance, velocity, and decrement in both distance and velocity over time. We considered only dyskinesia-free trials and only trials at stimulation frequencies 100 Hz or higher, whose half-harmonic was in the gamma range. In a linear mixed effects model, the presence of precentral gyrus gamma entrainment was associated with improved motor performance metrics (Fig. 4B) — specifically, greater maximum tap distance (*β* = 2.37, *p* < 0.001) and maximum tap velocity (*β* = 2.18, *p* = 0.004) — *after* controlling for DBS amplitude. DBS amplitude itself was associated with increased maximum tap distance (*β* = 0.56, *p* = 0.006). Notably, there was a negative interaction between DBS amplitude and gamma entrainment for both maximum tap distance (*β* = -0.81, *p* = 0.001) and maximum tap velocity (*β* = -0.71, *p* = 0.013), indicating a less-than-additive effect, consistent with a “ceiling” phenomenon with increasing amplitude (Fig. S3). Thus, cortical gamma entrainment contributes unique predictive value for clinical benefit beyond stimulation amplitude alone. The effect of STN gamma entrainment on finger tapping kinematics was less pronounced (Fig. S2B), although this may have been due to the lower detection rate for instances of STN gamma entrainment compared to the cortex.

For this finger-tapping kinematics analysis, we used the presence or absence of gamma entrainment during the rest epoch *immediately prior* to finger-tapping initiation to avoid the potential confound from the increase in gamma amplitude associated with movement. However, similar results were obtained when using the presence or absence of entrained gamma *during* the movement epoch (Fig. S4).

## Discussion

We studied the phenomenon of stimulation-entrained finely tuned gamma oscillations in a cohort of PD subjects previously implanted with a sensing-enabled pulse generator attached to subthalamic and sensorimotor cortical electrodes, in-clinic, in an ON-medication state. All patients had been on stable chronic stimulation. We utilized videotaped scoring of stimulation-induced motor benefits and adverse effects across a wide range of stimulation parameters. We found that stimulation frequencies between 90 and 150 Hz were the most efficient at producing half harmonic entrainment, that the amplitude of entrained oscillations was greater in the cortex than the STN, and that movement increased the amplitude of entrained cortical gamma oscillations. Stimulation parameters that entrained cortical gamma oscillations were associated with reduced risk of ON-period dyskinesia and dystonia, compared to parameters not associated with entrainment. The presence of cortical entrainment resulted in an improvement in finger-tapping kinematics beyond the effect of stimulation amplitude alone.

### Gamma oscillations in normal function and in PD

Gamma oscillations, first described in normal neocortical function, are a spectrally narrow form of oscillatory synchronization at frequencies above 30 Hz, arise from rhythmic interactions between excitatory and inhibitory interneurons^20,21^ and may coordinate interactions between cortical layers.^22^ These narrowband oscillations are distinct from broadband cortical activity, often referred to as “high gamma” or “broadband gamma”, an asynchronous process reflecting local cortical spiking activity.^23^ Within the motor system, the normal role of gamma oscillations has not been well studied, but transient bursts of gamma oscillations at 40-50 Hz occur in the motor cortex of nonhuman primates during movement preparation.^24^

In the human motor system, gamma oscillations at 60-90 Hz were first identified in the subthalamic nucleus in PD^4^. Narrowband activity in this specific frequency range has been called “finely tuned gamma”. Because this rhythm was induced by levodopa and anticorrelated with the “antikinetic” beta rhythm, STN FTG was considered a “prokinetic” rhythm. Subsequently, FTG was identified in the sensorimotor cortex in PD^8^ and found, in the off-stimulation condition, to predict risk for dyskinesia.^15^ Given the well described mechanism for the generation of gamma oscillations in the neocortex,^21^ it is possible that motor network FTG arises in the cortex and from there propagates to basal ganglia nuclei.

### Gamma entrainment and its effects on motor function

We first reported the phenomenon of half-harmonic entrainment of gamma oscillations in PD in an at-home study of motor cortex recordings in patients on chronic stimulation.^8^ Subsequently, across a larger cohort, we found entrained cortical gamma oscillations in 80% of subjects chronically stimulated at either STN or the globus pallidus.^12^ The conceptual framework for understanding entrainment phenomena is based on the physics of coupled oscillators, and neural entrainment has been modelled *in silico* using a simple neuronal network as the driven oscillator.^19^

Several recent studies evaluated entrained gamma activity in the subthalamic nucleus during chronic neurostimulation for PD. Mathiopoulou *et al*. found half harmonic entrainment of gamma oscillations in 15 of 19 subjects studied in-clinic.^9^ They proposed that entrained gamma is associated with improved motor outcomes since, across subjects, those with STN entrained activity had better motor outcomes. Two other studies identified entrained gamma in longitudinal recordings as the most reliable marker of the ON-medication state compared to oscillatory activity in other frequency bands.^10,11^ All of these studies found that *entrained* gamma activity is not consistently associated with a dyskinetic state, even though in prior human^15^ and animal^25,26^ studies, levodopa-induced gamma oscillations (in the *absence* of stimulation) are correlated strongly with risk of dyskinesia.

Here, we resolve this apparent paradox by demonstrating that stimulation induced entrainment of existing levodopa-associated gamma oscillations mitigates (though does not eliminate) the risk of on-period dyskinesia and dystonia. A similar entrainment-associated dyskinesia mitigation was found in chronic home recordings.^12^ Thus, not all gamma oscillations are alike; the behavioral effects of stimulation-entrained FTG differ from those of levodopa induced, non-entrained FTG. The mechanism by which entrainment mitigates dyskinesia risk is not certain, but it could involve the shift in frequency away from an individual’s “natural” gamma resonance frequency^12^ (the frequency associated with levodopa induced FTG). That is, the natural gamma frequency, abnormally “sharpened” by maladaptive cortico-striatal plasticity (due to chronic levodopa treatment)^27^ would confer dyskinesia risk, which is then mitigated by driving the system away from its zone of excessive resonance.

It remains unclear whether gamma entrainment at one site (basal ganglia or cortex) is required to produce entrainment at the other site. Here, subthalamic and cortical entrainment did not always co-occur. Entrained gamma activity measured in the STN likely arises from synapses *within* the STN (since it is these elements that generate the LFP), whereas long-distance entrainment of other structures might arise via stimulation of axons entering or exiting the STN^2^. Of note, pallidal stimulation is equally likely to produce cortical entrainment as subthalamic stimulation,^12^ but detection of entrained activity within the pallidum is uncommon, indicating that long-distance entrainment is dissociable from local entrainment. Given that the pallidum lacks a “hyperdirect” input from the cortex,^28^ cortical entrainment due to basal ganglia stimulation seems likely to occur via an orthodromic rather than an antidromic mechanism. The effects of therapeutic DBS on motor cortex are proposed to occur predominantly in the recently described somato-cognitive action network (SCAN) regions of motor cortex^29^ rather than the classical “effector” regions. We do not know if the quadripolar ECoG strips used in this study are indeed sampling SCAN zones preferentially, but their placement 2-4 cm from midline does correspond to the most mesial of the three SCAN zones,^30^ raising the possibility that gamma entrainment occurs preferentially in the SCAN.

The finding that the presence of gamma entrainment is associated with improved bradykinesia in the ON-medication state (while controlling for stimulation amplitude) suggests that the prokinetic benefits for medication and stimulation therapy are complementary rather than redundant. While clinicians tend to counsel patients that optimized stimulation only achieves the same “maximum” benefit as the best ON-medication state,^31^ our results suggest that in fact, full engagement of the motor network as evidenced by cortical entrainment confers an additive benefit in bradykinesia, even during ON-medication states.

### A new paradigm for physiologically-guided simulation programming

Most prior studies of physiology-guided programming have focused on identifying the basal ganglia contact with the strongest “antikinetic” beta activity, in the OFF-medication state, and increasing stimulation until beta is suppressed.^32,33^ However, full beta suppression often occurs at a low-therapeutic amplitude rather than full-therapeutic amplitude.^11,33^ We propose that the full therapeutic amplitude might be determined by the optimization of stimulation entrained gamma activity. A gamma-based programming paradigm would proceed by establishing the boundaries of entrainment in the frequency-amplitude space by direct measurement or by modeling approaches^19^. Stimulation settings which optimize entrainment would also maximize motor improvement and mitigate dyskinesia risk. A high degree of entrainment does not necessarily correspond to the highest tolerated amplitude, as increasing amplitude may be associated with loss of half harmonic entrainment. Since entrainment is established within seconds of changing stimulation settings, this programming method could be automated and accomplished quickly.

Since cortical electrodes are not routinely implanted for surgical therapy it would be ideal if subthalamic entrainment could be used exclusively for evaluation of gamma entrainment. In general, subthalamic entrainment showed similar predictive trends as cortical entrainment in this data set, but of lesser magnitude and significance. This may be related to the lower detection rate for STN entrainment. There are advantages to cortical detection: it is larger in amplitude (using invasive recording) and is more readily dissociated from stimulation artifact, especially when using stimulation contacts that are not “sense friendly” for basal ganglia local field potential recording. Cortical entrainment also reflects long-distance circuit engagement across the motor network which may be mechanistically important for providing additional therapeutic benefit and/or mitigating hyperkinetic states. Scalp EEG could provide a noninvasive alternative for cortical detection.^5^

The proposed use of entrainment phenomena in clinical programming for PD need not be restricted to persons with pre-existing levodopa-generated gamma oscillations. Chronic stimulation can produce entrained gamma oscillations even in subjects in whom, off stimulation and on medication, levodopa-associated gamma oscillations were not detected.^9,12^ Thus, therapeutic stimulation can both induce *and* entrain gamma oscillations. Further, entrainment-based programming might be applicable to other movement disorders. Cortical^34^ and subthalamic^35^ FTG are also features of isolated dystonia, and entrainment of motor cortical gamma activity by pallidal stimulation indexes therapeutic benefit for this disorder as well.^13^

### Limitations

Study results may have been affected by prior chronic stimulation, biasing the efficiency of entrainment toward the clinically utilized frequency and contact choice via plasticity mechanisms. Detection of subthalamic entrainment might be further enhanced by higher spatial sampling, especially with directional leads, which were not used here. We had insufficient data to compare effects of entrainment at the “natural” levodopa induced gamma frequency, versus entrainment that shifts gamma activity away from its “natural” frequency.

### Conclusion

Entrainment of gamma oscillations by therapeutic neurostimulation occurs most efficiently at stimulation frequencies of 90-150 Hz, which correspond to the classic frequency range empirically determined to be efficacious since the introduction of DBS for PD. Entrained gamma oscillations were associated with beneficial effects on motor function that are distinct from those of levodopa-induced, un-entrained gamma oscillations, and could prove useful in guiding DBS programming.

## Data Availability

All data produced in the present study are available upon reasonable request to the authors.

## Acknowledgments

The authors are appreciative of the patients who participated in the study. The study was supported by National Institute of Neurological Disorders and Stroke (NINDS) UH3NS100544. Investigational devices were provided at no charge by the manufacturer, but the manufacturer had no role in the conduct, analysis or interpretation of the study. The Open Mind consortium for technology dissemination provided technical resources for the use of the Summit RC+S neural interface. The authors thank Colin W. Hoy for assistance with figure preparation.

## Author’s Roles

MS, SC and PAS designed the experiments. MS, SC, and AGH executed the experiments. MS performed the analysis. MS wrote the manuscript. MS, SC, AGH, SL and PAH edited the final version of the manuscript.

