## Supplementary material for "Entrainment of cortical gamma oscillations predicts improved bradykinesia and dyskinesia in Parkinson’s disease": Figure S1, Figure S2, Figure S3, Figure S4, Supplementary Methods

Shcherbakova M. DBS-entrained gamma and motor improvement

**SUPPLEMENTARY MATERIALS**

### Supplemental Figures

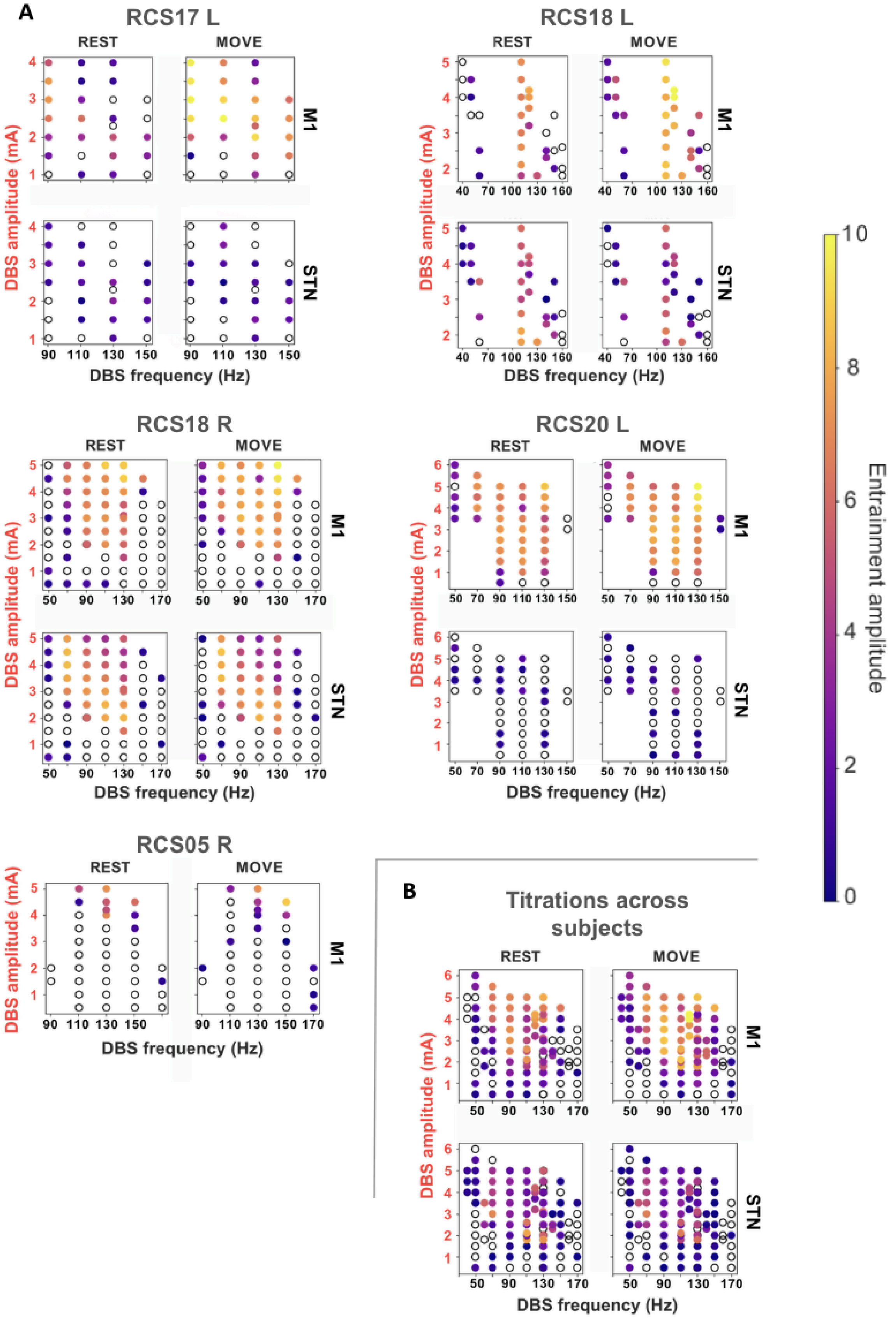

**Figure S1. Amplitude of entrained oscillations as a function of stimulation frequency and amplitude, at the clinically chosen stimulation contact,** plotted for each hemisphere individually (**A**) and across all patients (**B**). In (**B**), amplitudes for trials testing the same frequency-amplitude settings were averaged. *(Note: S1 showed patterns similar to M1 and is omitted for simplicity)*. The exact parameter combinations that result in entrainment in STN and motor cortex differ within and across subjects.

**A** Co-occurrence of STN entrainment and motor signs across patients

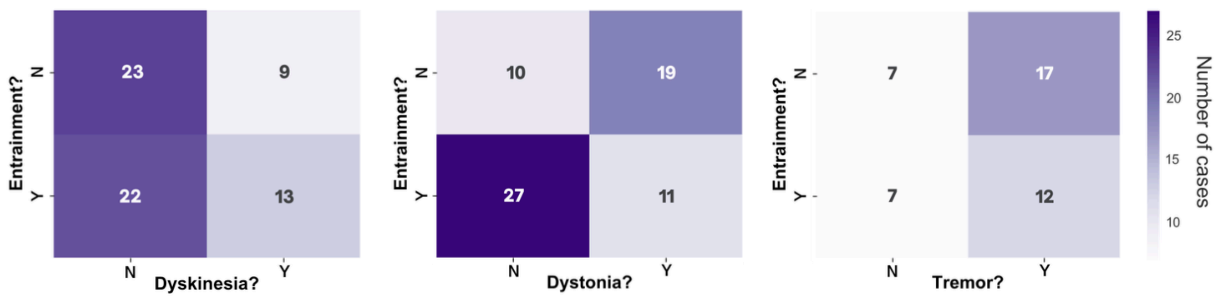


**B**

| Outcome metric | Gamma entrainment (Y/N) Coef. ($\beta$) | Gamma entrainment (Y/N) $p$-value | Stimulation amplitude, mA Coef. ($\beta$) | Stimulation amplitude, mA $p$-value | Interaction, gamma:stimulation Coef. ($\beta$) | Interaction, gamma:stimulation $p$-value |
|---|---|---|---|---|---|---|
| AVG tap distance | 0.678 | 0.324 | 0.164 | 0.317 | -0.221 | 0.342 |
| MAX tap distance | 1.061 | 0.09 | 0.265 | 0.081 | -0.393 | 0.07 |
| Decrement, distance | N/A | N/A | N/A | N/A | N/A | N/A |
| AVG tap velocity | 0.731 | 0.282 | 0.13 | 0.424 | -0.287 | 0.214 |
| MAX tap velocity | 1.689 | **0.014** | 0.234 | 0.168 | -0.611 | **0.012** |
| Decrement, velocity | -0.225 | 0.713 | -0.351 | **0.016** | 0.312 | 0.131 |

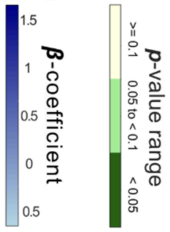


**Figure S2. Effects of STN gamma entrainment on motor function. A**: 2x2 contingency tables showing the association between STN gamma entrainment and common parkinsonian motor signs observed in clinic. *Note that this is a more limited data set than that shown in Figure 4A since subject RCS05 only contributed cortical data to the study.* **B:** Summary of the results of a linear mixed effects model to quantify the effects of gamma entrainment at *therapeutic* stimulation frequencies (≥100 Hz) on predefined metrics of bradykinesia obtained from video kinematics. As predictors, the model uses 1) presence of gamma entrainment, recorded immediately *prior* to movement onset (finger-tapping task), and 2) stimulation amplitude. *Coefficients show the direction and magnitude of relationships, with greater absolute β-values signifying stronger association.*

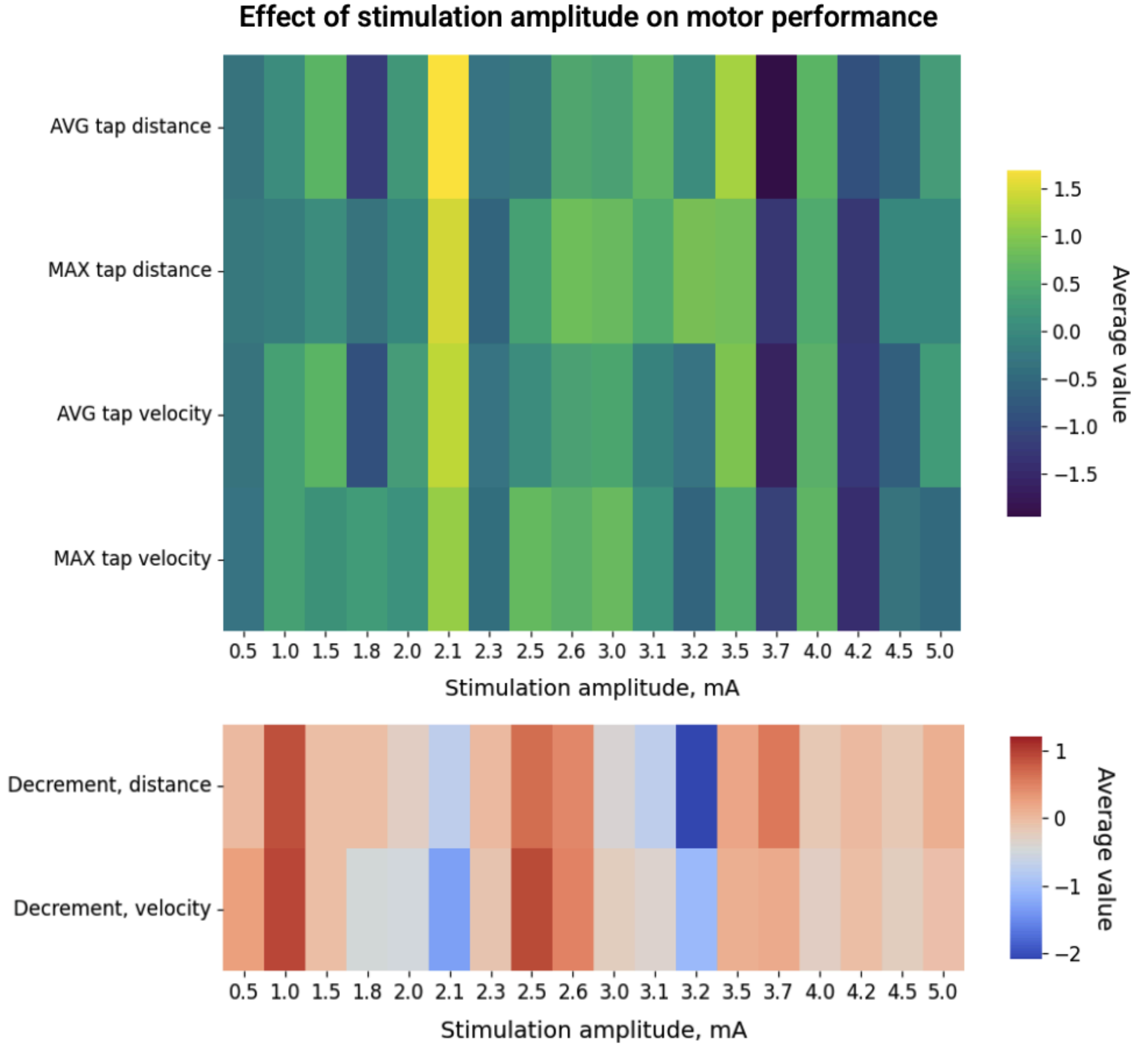


**Figure S3. Ceiling effect of stimulation on improvement of movement kinematics.** Increasing DBS amplitude alone is not associated linearly with improving finger-tapping task performance. ***Top*** *(indigo-yellow)*: higher kinematic values indicate improvement; ***bottom*** *(blue-red)*: lower kinematic values indicate improvement.

| | Gamma entrainment (Y/N) | | Stimulation amplitude, mA | | Interaction, gamma:stimulation | |
|---|---|---|---|---|---|---|
| Outcome metric | Coef. ($\beta$) | $p$-value | Coef. ($\beta$) | $p$-value | Coef. ($\beta$) | $p$-value |
| AVG tap distance | 1.036 | 0.195 | 0.214 | 0.46 | -0.301 | 0.362 |
| MAX tap distance | 2.198 | **0.002** | 0.434 | 0.082 | -0.681 | **0.018** |
| Decrement, distance | -1.708 | **0.02** | -0.587 | **0.032** | 0.716 | **0.019** |
| AVG tap velocity | 0.951 | 0.227 | 0.11 | 0.702 | -0.268 | 0.414 |
| MAX tap velocity | 2.19 | **0.006** | 0.295 | 0.31 | -0.676 | **0.04** |
| Decrement, velocity | -1.382 | 0.068 | -0.42 | 0.12 | 0.484 | 0.111 |

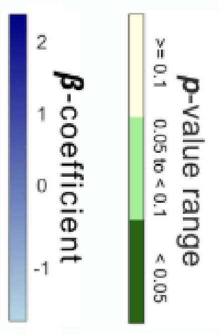

**Figure S4. Effects of motor cortical gamma entrainment on motor function: repeat of linear mixed model.** In contrast to Figure 4B, this model relies on the presence of gamma entrainment using signals recorded *during* ongoing movement rather than those recorded *before*.

### Supplementary Methods

Further details on selection of study subjects and laterality of recordings

Individuals were selected for the present study, from the larger parent study of chronic multisite neural recording in Parkinson's disease, based on 2 criteria: 1) willingness to travel to the research site at the University of California, San Francisco for additional in-person studies, and 2) prior demonstration of ability to detect cortical and STN entrained gamma oscillations.[11,12] Of note, in one subject (RCS05), study of STN entrainment was precluded by the occurrence of stimulation artifacts of unclear origin.

For the three subjects recorded unilaterally, the hemisphere selected for study was the one previously known to exhibit either STN or cortical entrained gamma at clinical stimulation settings. It was also the hemisphere contralateral to the side of the body with more severe motor signs. The subject studied on both hemispheres (RCS18) underwent recording in two different sessions separated by 2 months, and had severe motor signs on both sides. Reduction in levodopa had been done between recordings in order to allow an increase in right sided stimulation.

Details of hand tracking analysis

Video input was first pre-processed using OpenCV.[36] Palm detection and motion capture were performed using the Hand Landmarker python library by Google AI,[37,38] a high-fidelity machine-learning framework for hand and finger tracking. Please see referenced materials for detailed installation instructions.
